# Trends, Rural–Urban Inequalities and Forecasts of Open Defecation in Ghana Using Joinpoint Regression and ARIMA Analysis, 2000–2030

**DOI:** 10.64898/2026.06.24.26356484

**Authors:** Abdul-Wahab Inusah, Moses Ifeatu Nwuzoh, Abdul-Aziz Seidu

## Abstract

**Background:** Open defecation remains a major public health challenge in Ghana and across sub-Saharan Africa, with persistent rural–urban inequalities undermining progress toward Sustainable Development Goal 6.2 (SDG 6.2). Despite two decades of national sanitation programming, structural and equity barriers continue to constrain progress. A repeated cross-sectional analyses combining WHO-standardised inequality measures, temporal trend modelling, and projections remain absent from the literature for Ghana.

**Methods:** National, rural, and urban open defecation prevalence (2000–2024) was analysed using WHO Health Equity Assessment Toolkit (HEAT) data. Four inequality measures: Difference, Ratio, Population Attributable Risk (PAR), and Population Attributable Fraction (PAF), quantified rural–urban disparities. Joinpoint regression identified statistically significant trend inflection points across MDG and SDG eras. ARIMA models projected prevalence to 2030 under status quo, accelerated, and decelerated scenarios; hold-out validation confirmed high forecast accuracy across all series (MAPE <1%).

**Results:** National prevalence declined from 20.31% to 17.79% (AAPC: −0.55%, p<0.001), with a joinpoint at 2016 (95% CI: 2015–2017) after which decline slowed during the SDG era. Rural prevalence rose marginally (AAPC: +0.07%) with no significant joinpoints across the 25-year period; urban prevalence also increased (AAPC: +0.76%). Rural prevalence exceeded urban more than threefold by 2024 (R=3.38); PAF improved from −62.62% to −48.85%, indicating a substantial national burden attributable to rural disadvantage. Under the status quo scenario, national and rural prevalence are projected at 17.24% and 30.88% by 2030, far exceeding the SDG 6.2 threshold.

**Conclusion:** Despite modest national progress, substantial rural–urban inequalities remain entrenched, and Ghana is unlikely to achieve SDG 6.2 under current trajectories. Accelerated, equity-focused interventions targeting structurally disadvantaged rural populations are urgently required to reduce sanitation inequalities and improve health outcomes.

## Background

Access to safe sanitation is a fundamental determinant of human health, dignity, and development. Sustainable Development Goal 6.2 (SDG 6.2) commits countries to achieving universal access to adequate and equitable sanitation and hygiene and eliminating open defecation by 2030, with particular attention to women, girls, and vulnerable populations [1–3]. Open defecation, defined as defecating in fields, forests, water bodies, or other open spaces rather than using toilets or latrines, remains a major and preventable public health challenge. It contributes to diarrhoeal diseases, cholera, typhoid, and soil-transmitted helminthiases, undermines child nutrition and cognitive development, and perpetuates poverty, particularly among women and girls who face heightened risks of gender-based violence and educational disruption [4–6].

Globally, an estimated 419 million people practised open defecation in 2022, with sub-Saharan Africa bearing a disproportionate share of the burden as population growth continues to outpace sanitation infrastructure expansion [7]. A spatial analysis across 34 sub-Saharan African countries estimated regional open defecation prevalence at 23.24% (95% CI: 23.12–23.35), with marked clustering across East, Central, and West Africa [8]. Current trends indicate that many countries remain off track toward SDG 6.2, with open defecation declining by only 0.22% annually between 2015 and 2020 [9]. Scenario modelling further projects that universal improved sanitation may not be achieved before 2070–2090 under optimistic pathways and beyond 2100 under pessimistic scenarios [10]. Low-and lower-middle-income countries are estimated to require a six-to tenfold acceleration in current progress rates to eliminate open defecation by 2030 [11].

Within this regional context, Ghana represents an important case study. Despite sustained macroeconomic growth over the past two decades, the country continues to experience persistent sanitation deficits [12]. National estimates conceal substantial subnational inequalities: coastal urban zones report relatively low open defecation prevalence (6.7%), whereas Northern and Middle zones report rates of 15.2% and 12.5%, respectively [13]. Among rural women aged 15–49 years, approximately 42% practise open defecation [12]. Ghana also has one of the highest proportions of open-pit latrine use in sub-Saharan Africa, estimated at approximately 50% and remaining relatively stable between 2001 and 2017 [14]. Repeated cross sectional analysis surveillance from the Kintampo Health and Demographic Surveillance System further reported that 44.2% of households practised open defecation in 2016, with projections suggesting Ghana would not eliminate open defecation by 2030 under prevailing trajectories [15].

The rural–urban divide in sanitation extends beyond differences in service coverage and reflects broader socioeconomic and geographic inequalities. Across sub-Saharan Africa, rural residents are estimated to be three to ten times more likely to practise open defecation than urban populations [16]. In Ghana, poverty, lower educational attainment, male household headship, younger age, northern residence, and limited media exposure are consistently associated with elevated open defecation prevalence [12,17]. Residents of Savannah ecological zones demonstrate particularly high risk, highlighting the compounding effects of geography and deprivation [12].

Ghana formally adopted the Community-Led Total Sanitation (CLTS) approach in 2008, following its pioneering development in Bangladesh and subsequent diffusion across sub-Saharan Africa. CLTS employs facilitated community-level participatory analysis of local sanitation practices to trigger collective behaviour change and motivate open defecation-free (ODF) commitments without subsidising household latrine construction [18]. The approach was scaled nationally through the Community Water and Sanitation Agency (CWSA) and was embedded within Ghana’s 2010 National Environmental Sanitation Policy and the subsequent Environmental Sanitation Policy Action Plan. By 2014, CLTS had been implemented across all the then 10 regions, with ODF verification processes progressively integrated into district-level sanitation programming [19]. Despite early gains in triggering community engagement, sustaining ODF status has remained a persistent challenge, with evidence of sanitation slippage, the reversion to open defecation following initial ODF certification, documented in multiple districts, particularly in rural Northern Ghana [20]. Although these determinants are increasingly recognised, repeated cross sectional quantification of rural–urban inequalities in Ghana using internationally standardised summary measures, including Difference (D), Ratio (R), Population Attributable Risk (PAR), and Population Attributable Fraction (PAF), remains absent from the literature.

Understanding how sanitation trends evolve over time is also critical for evaluating the effectiveness of policy responses. Joinpoint regression has become a widely validated approach for identifying statistically significant changes in temporal health trends, including accelerations, decelerations, and reversals [21,22]. Although commonly applied in cancer epidemiology, non-communicable disease surveillance, and maternal and child health research, its application to sanitation trends in Ghana and sub-Saharan Africa remains limited. Examining not only whether open defecation prevalence changes over time, but also when significant shifts occur, particularly across the transition from the Millennium Development Goals (MDGs, 2000–2015) to the SDGs (2016–2030), is important for assessing the impact of sanitation policies and programme implementation.

Forecast analysis is similarly important for translating epidemiological trends into policy-relevant evidence. While global modelling studies have projected sanitation trajectories [9,10], Ghana-specific forecasts using disaggregated rural–urban data and incorporating the most recent surveillance estimates through 2024 remain lacking. The WHO Health Equity Assessment Toolkit (HEAT) provides a standardised platform for such analyses, enabling nationally representative and equity-disaggregated assessment of open defecation trends over time [23].

Against this backdrop, the present study addresses three interconnected objectives: (1) to quantify the magnitude and temporal evolution of rural–urban inequalities in open defecation using WHO standardised inequality measures; (2) to characterise national and residence-specific trends in open defecation prevalence in Ghana from 2000 to 2024 using joinpoint regression; and (3) to generate forecasts of Ghana’s open defecation trajectory through 2030 to assess progress toward SDG 6.2.

This study contributes to the literature in three important ways. First, it integrates four complementary WHO-standardised inequality measures (D, R, PAR, and PAF) within a unified framework to provide a multidimensional assessment of sanitation inequality. Second, it applies joinpoint regression to a 25-year sanitation time series in Ghana, enabling identification of trend inflection points across major policy periods, including the MDG-to-SDG transition and the implementation of Community-Led Total Sanitation initiatives. Third, it provides updated Ghana-specific forecasts through 2030 using data up to 2024, generating policy-relevant evidence on the country’s likely SDG 6.2 trajectory. These contributions provide a comprehensive analytical framework for understanding sanitation inequalities and trajectories in Ghana and offer a replicable approach for similar analyses across sub-Saharan Africa.

## Methods

### Study Design and Setting

This study employed a secondary data analysis design using repeated cross-sectional survey data spanning 25 years (2000–2024). The study setting is the Republic of Ghana, a West African lower-middle-income country with an estimated population of approximately 34 million in 2024 [24]. Ghana is administratively organised into 16 regions characterised by marked ecological, economic, and infrastructural heterogeneity, which is reflected in substantial subnational variation in sanitation outcomes. The study period spans both the MDG era (2000–2015) and the early SDG era (2016–2024), enabling direct comparative assessment of trend trajectories across two major global development policy frameworks.

### Data Source

Data were obtained from the World Health Organization Health Equity Assessment Toolkit (WHO HEAT) platform (version 4.0), an open-access database providing standardised, equity-disaggregated estimates of health indicators across multiple dimensions of inequality for WHO Member States [25]. The HEAT platform draws on data from nationally representative household surveys, including Demographic and Health Surveys (DHS) and Multiple Indicator Cluster Surveys (MICS), as well as WHO/UNICEF Joint Monitoring Programme (JMP) estimates, processed through standardised methodological protocols to ensure temporal and cross-national comparability.

The indicator of interest was Population practising open defecation (%), defined as the percentage of the population using no sanitation facility and defecating in fields, forests, bodies of water, or other open spaces, consistent with WHO/UNICEF JMP definitions and measurement standards [26]. Data were extracted for Ghana for the years 2000 through 2024, disaggregated by place of residence (rural and urban), yielding a 25-observation national time series and two parallel 25-observation subgroup series. No primary data collection was undertaken, and no ethical approval was required under institutional guidelines for the analysis of publicly available, de-identified, aggregated datasets. Data access and utilisation complied fully with the WHO HEAT platform’s terms of use.

### Variables

The primary outcome variable was open defecation prevalence (%), measured at the national, rural, and urban levels for each year from 2000 to 2024. The primary disaggregation variable was place of residence (rural vs. urban), which served as the equity stratifier for inequality analysis.

### Analytical Approach

Descriptive analysis was conducted as the foundational step to characterise the distribution and temporal patterns of open defecation prevalence in Ghana across the 25-year study period. National open defecation prevalence estimates were tabulated annually from 2000 to 2024 alongside rural and urban subgroup estimates, enabling visual and numerical assessment of overall burden, residence-based differentials, and the direction of change over time. Trends were initially examined through graphical plots of annual prevalence series for national, rural, and urban estimates, providing an exploratory basis for the subsequent analytical modelling. All prevalence estimates are reported as percentages.

The study then employed three complementary analytical methods, each addressing a distinct but interconnected research objective, selected to provide both epidemiological rigour and policy relevance. Summary measures of rural–urban inequality were computed directly within the WHO HEAT platform (version 4.0), which provides an integrated analytical environment for generating standardised inequality metrics across disaggregated population subgroups [25]. Joinpoint regression analysis was conducted using the Joinpoint Regression Program (version 6.0.1) to characterise national and subgroup-specific open defecation trends and identify statistically significant inflection points over the 25-year study period [27]. Forecast modelling extending observed trends through 2030 was performed in Python (version 3.11) [28], enabling scenario-based assessment of Ghana’s likelihood of achieving SDG 6.2. Supporting descriptive tabulations and prevalence computations were undertaken in Microsoft Excel 2021. A two-tailed significance threshold of α = 0.05 was applied uniformly across all inferential procedures.

### Inequality Analysis: WHO Summary Measures

To quantify the magnitude and temporal evolution of rural–urban inequalities in open defecation prevalence, four WHO-recommended summary measures of health inequality were computed for each year from 2000 to 2024 directly within the WHO Health Equity Assessment Toolkit (HEAT) platform (version 4.0), following established WHO HEAT methodological guidelines [22]. These measures collectively capture both the absolute and relative dimensions of inequality, providing a comprehensive multi-dimensional characterisation of equity dynamics that is consistent with WHO best-practice guidance and internationally comparable across settings [22,29]. Place of residence (rural vs. urban) served as the equity stratifier, with urban residents designated as the reference (most advantaged) group in all attributable risk computations.

The four summary measures were defined and interpreted as follows:

### Absolute Difference (D)

The simple difference in open defecation prevalence between rural and urban subgroups, expressed in percentage points:

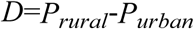

A positive value indicates higher prevalence in rural areas relative to urban areas. D directly quantifies the absolute equity gap and is interpretable in policy terms as the magnitude of the rural disadvantage to be eliminated. It is sensitive to changes in both subgroup rates and is directly actionable for programme targeting.

### Rate Ratio (R)

The ratio of rural open defecation prevalence to urban open defecation prevalence:

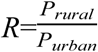

R is a relative measure of inequality that is independent of the absolute level of prevalence, enabling meaningful comparison of proportional disparities across time points and country settings. A value greater than 1.0 indicates rural disadvantage; a value of 1.0 indicates perfect parity between residence groups. R is particularly informative in settings where absolute prevalence is declining, as it captures whether relative disparities are narrowing or widening independently of overall progress.

### Population Attributable Risk (PAR)

The difference between the observed national open defecation prevalence and the prevalence that would be expected if all population subgroups experienced the same rate as the reference (most advantaged) group, in this case, urban residents:

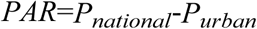

PAR is expressed in percentage points and quantifies the excess national burden of open defecation attributable to the rural–urban disparity. It estimates the potential reduction in national prevalence that would be achievable if rural open defecation were reduced to the level observed in urban areas, providing a direct benchmark for equity-driven policy targets.

### Population Attributable Fraction (PAF)

The proportion of total national open defecation prevalence attributable to the rural–urban inequality, expressed as a percentage:

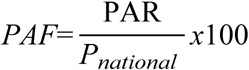

PAF indicates the fraction of the national open defecation burden that could theoretically be eliminated by reducing rural prevalence to the urban reference level. Unlike PAR, PAF is a relative measure that adjusts for the overall national burden, making it particularly useful for tracking the proportional contribution of the rural–urban divide to national open defecation over time and for comparing the equity dimension of the burden across different periods.

### Joinpoint Regression Analysis

Joinpoint regression analysis was employed to detect statistically significant changes in the annual prevalence of open defecation in Ghana from 2000 to 2024, disaggregated by national estimates and place of residence (rural and urban). This model identifies points in time, referred to as joinpoints, at which statistically significant changes occur in the direction or magnitude of the trend, and provides confidence measures around these estimated changes. The analytical approach enables precise characterisation of whether open defecation prevalence accelerated, decelerated, or reversed direction at specific points during the study period, including across the transition from the MDG era (2000–2015) to the SDG era (2016–2024). All analyses were conducted using the Joinpoint Regression Program, Version 6.0.1 [27].

The Joinpoint program fits a series of joined straight lines to the open defecation prevalence estimates on a logarithmic scale, identifying the best-fitting inflection points that represent statistically significant changes in trend [21]. Model selection was primarily based on the Monte Carlo permutation method implemented in the Joinpoint Regression Program, while the Bayesian Information Criterion (BIC) was used as a complementary goodness-of-fit indicator. The analysis begins with the minimum number of joinpoints, zero joinpoints, equivalent to a single straight line representing a constant rate of change, and tests sequentially whether the addition of one or more joinpoints significantly improves model fit. Based on the recommendation of the Joinpoint program and consistent with the 25 data points available in this study, a maximum of four joinpoints was permitted, following established guidance that the maximum number of joinpoints should not exceed the number that would result in fewer than four observations per segment, and the minimum jointpoints was zero, these were applied consistently for national, rural and urban [22,30].

The permutation test determines the optimal number of joinpoints through a sequential hypothesis-testing procedure. Beginning with a minimum number of joinpoints k_a_ and a maximum k_b_, each test compares the null hypothesis H_0_: number of joinpoints = k_a_ against the alternative hypothesis H_a_: number of joinpoints = k_b_, where k_a_ < k_b_. If the null hypothesis is rejected, k_a_ is increased by one; otherwise k_b_ is decreased by one. This process continues iteratively until k_a_ = k_b_, at which point the final number of joinpoints is determined. Models with this optimal number of joinpoints are then compared using BIC, and the model yielding the minimum BIC value is selected as the final optimal model [21,30].

To quantify the magnitude and direction of change in open defecation prevalence within each identified trend segment and across the full study period, two summary statistics were computed with corresponding 95% confidence intervals (95% CI): the Annual Percentage Change (APC) and the Average Annual Percentage Change (AAPC). Log transformation was applied prior to regression modelling, consistent with the assumption that open defecation prevalence changes at a constant percentage relative to the prevalence of the preceding year, thereby allowing linear modelling on a logarithmic scale. The underlying regression model fitted to the open defecation prevalence series is expressed as:

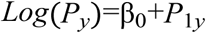

where log (P_y_) is the natural logarithm of the open defecation prevalence in year, y, β_0_ is the intercept representing the log prevalence when y=0, and β_1_ is the slope coefficient indicating the annual rate of change in log prevalence. The APC from year y to year y+1 is derived as:

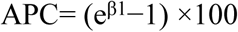

The AAPC summarises the overall trend across the entire study period (2000–2024) as a single weighted average of the segment-specific APCs, with weights proportional to the length of each joinpoint segment within the period of interest:

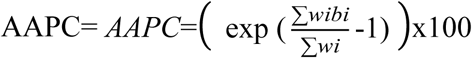

Where b_i_ is the slope coefficient for the *i*th segment and w_i_ is the length of that segment in years. The AAPC thus provides a single summary measure of the average annual rate of change in open defecation prevalence over the full 25-year period, accounting for the varying durations and slopes of each trend segment identified by the joinpoint model.

Joinpoint regression was performed separately for the national open defecation series and for the rural and urban residence subgroups, enabling direct comparison of trend trajectories and inflection point timing across the equity dimension of interest. A trend was classified as statistically significantly decreasing when the APC was negative and the corresponding p-value was below 0.05, statistically significantly increasing when the APC was positive and p < 0.05, and non-significant when p ≥ 0.05. This classification framework provides a rigorous basis for characterising whether Ghana’s open defecation reduction has been sustained, accelerating, or stalling within each sub-period of the MDG and SDG eras.

### Forecast Analysis and Projections to 2030

To assess Ghana’s projected trajectory toward the SDG 6.2 target of ending open defecation by 2030, forecast modelling was conducted using observed open defecation prevalence data from 2000 to 2024 for national, rural, and urban series. All analyses were performed in Python (version 3.11; Python Software Foundation) using the *statsmodels* library [28].

An Autoregressive Integrated Moving Average (ARIMA) framework was employed as the primary forecasting method, given its suitability for univariate time series data with temporal autocorrelation and non-stationarity [31]. Prior to model fitting, stationarity of each series was assessed using the Augmented Dickey-Fuller (ADF) test, with first-order differencing applied where non-stationarity was detected. Optimal model order parameters (p, d, q) were selected using the Akaike Information Criterion (AIC), and residual adequacy was confirmed using the Ljung-Box test. Forecasts with 95% prediction intervals were generated for the period 2025 to 2030 for national, rural, and urban series.

Model performance was evaluated through a hold-out validation approach, fitting the model to data from 2000 to 2019 and assessing accuracy against observed values from 2020 to 2024. Three metrics were computed: Mean Absolute Error (MAE), Root Mean Square Error (RMSE), and Mean Absolute Percentage Error (MAPE). A MAPE below 10% was considered indicative of high forecast accuracy [32].

Projected 2030 prevalence estimates were benchmarked against the SDG 6.2 near-elimination threshold of less than 1% open defecation prevalence, consistent with WHO/UNICEF Joint Monitoring Programme definitions [7,26]. To bound the range of plausible outcomes and inform scenario-based policy planning, three projection scenarios were constructed: a status quo scenario maintaining the most recently observed rate of change; an accelerated scenario incorporating a 50% amplification of the absolute annual rate of change, representing the potential impact of intensified programmatic investment and equity-focused rural targeting where declining series fall faster and rising series rise faster; and a deceleration scenario incorporating a 50% dampening of the absolute annual rate of change, reflecting potential programmatic stagnation or emerging structural barriers where declining series fall more slowly and rising series rise more slowly. It should be noted that for the rural and urban series, which exhibited upward trends across the study period, the accelerated scenario projects faster increases in prevalence and the deceleration scenario projects slower increases, as the scenario logic amplifies or dampens the direction of the fitted trend regardless of its sign. The absolute gap between projected 2030 prevalence and the SDG 6.2 near-elimination threshold of less than 1% open defecation prevalence was quantified for each scenario and residence subgroup to identify populations at greatest risk of being left behind.

## Results

Open defecation in Ghana showed a gradual decline at the national level from 20.31% in 2000 to 17.79% in 2024. This period spans the MDG and SDG eras, covering two distinct global development frameworks for sanitation improvement. Despite this long-term policy transition, the overall reduction remained modest over time.

At the subnational level, divergent patterns were observed. Rural open defecation remained persistently high throughout the period, increasing marginally from 30.28% to 30.76%, indicating near-stable levels with minimal change over time. In contrast, urban prevalence increased from 7.59% to 9.10%, reflecting a slow upward trajectory across the same period. These contrasting trends resulted in a persistent rural–urban inequality throughout the study period. The absolute gap narrowed slightly from 22.69 percentage points in 2000 to 21.66 percentage points in 2024, indicating very limited convergence between rural and urban populations over the 24-year period. Annual estimates of national, rural, and urban prevalence, along with the rural–urban gap, are summarised in Table 1.

**Table 1:**
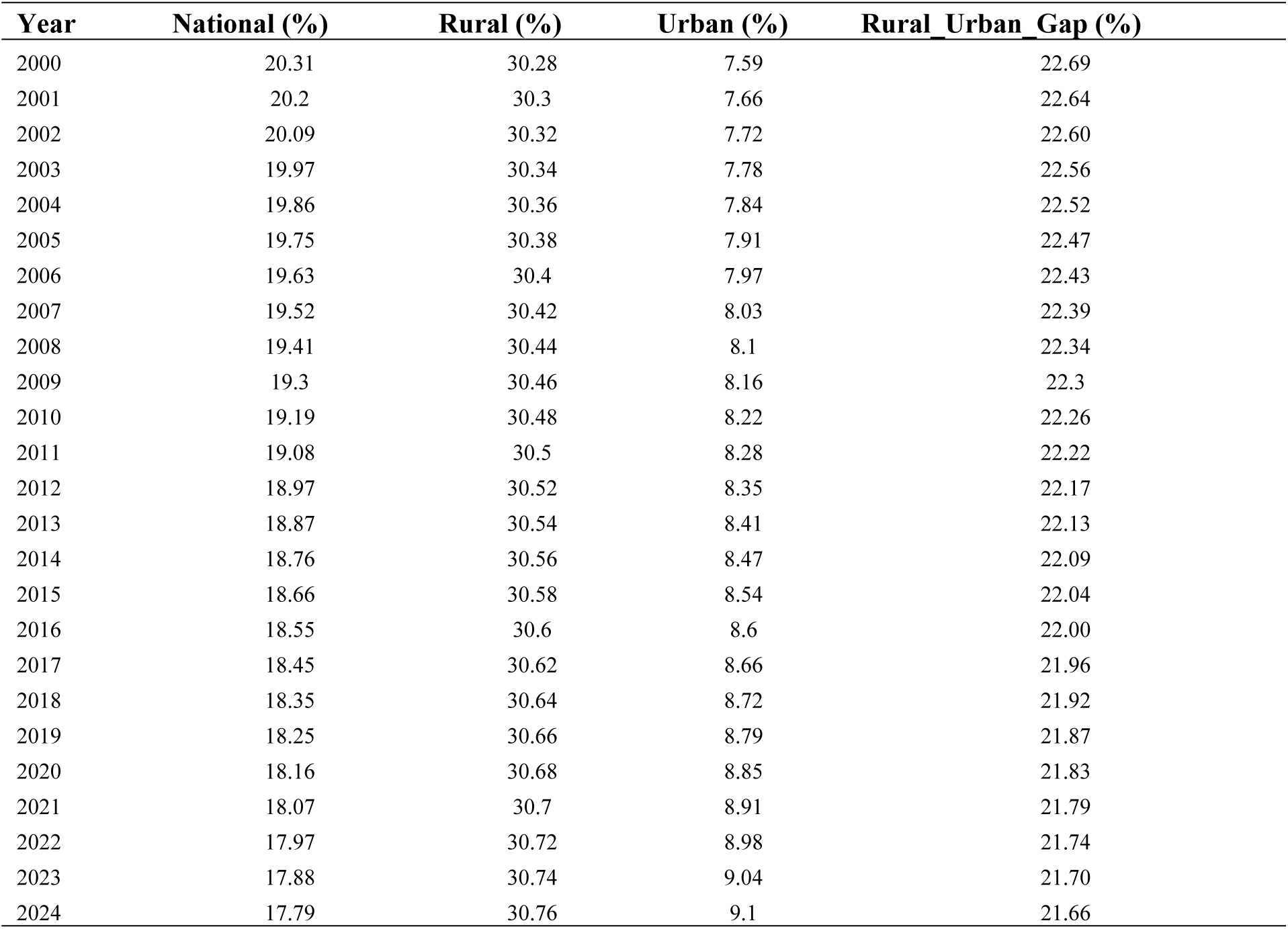
National, rural, and urban open defecation prevalence and rural–urban gap in Ghana, 2000–2024.

### Summary measures

As shown in Table 2, rural–urban inequality in open defecation persisted throughout the 2000–2024 period, with only gradual and limited convergence between population groups. The absolute difference (D) declined modestly from 22.68 percentage points in 2000 to 21.65 percentage points in 2024, indicating a slow narrowing of the rural–urban gap over time. The decline was gradual and relatively stable across the study period, with only minor year-to-year variation.

**Table 2:** Trends in rural–urban inequality in open defecation in Ghana, 2000–2024: summary measures of absolute and relative disparity.

| Year | D | R | PAF | PAR |
| --- | --- | --- | --- | --- |
| 2000 | 22.68 | 3.99 | -62.62 | -12.72 |
| 2001 | 22.64 | 3.96 | -62.10 | -12.54 |
| 2002 | 22.60 | 3.93 | -61.57 | -12.37 |
| 2003 | 22.55 | 3.90 | -61.04 | -12.19 |
| 2004 | 22.51 | 3.87 | -60.50 | -12.01 |
| 2005 | 22.47 | 3.84 | -59.96 | -11.84 |
| 2006 | 22.43 | 3.81 | -59.41 | -11.66 |
| 2007 | 22.38 | 3.79 | -58.85 | -11.49 |
| 2008 | 22.34 | 3.76 | -58.29 | -11.32 |
| 2009 | 22.30 | 3.73 | -57.73 | -11.14 |
| 2010 | 22.25 | 3.71 | -57.16 | -10.97 |
| 2011 | 22.21 | 3.68 | -56.58 | -10.80 |
| 2012 | 22.17 | 3.66 | -56.00 | -10.62 |
| 2013 | 22.13 | 3.63 | -55.42 | -10.45 |
| 2014 | 22.08 | 3.61 | -54.83 | -10.29 |
| 2015 | 22.04 | 3.58 | -54.24 | -10.12 |
| 2016 | 22.00 | 3.56 | -53.65 | -9.95 |
| 2017 | 21.95 | 3.53 | -53.06 | -9.79 |
| 2018 | 21.91 | 3.51 | -52.46 | -9.63 |
| 2019 | 21.87 | 3.49 | -51.86 | -9.47 |
| 2020 | 21.83 | 3.47 | -51.26 | -9.31 |
| 2021 | 21.78 | 3.44 | -50.66 | -9.15 |
| 2022 | 21.74 | 3.42 | -50.06 | -9.00 |
| 2023 | 21.70 | 3.40 | -49.45 | -8.84 |
| 2024 | 21.65 | 3.38 | -48.85 | -8.69 |

A similar pattern was observed for the relative inequality measure. The ratio (R) decreased from 3.99 in 2000 to 3.38 in 2024, indicating that rural open defecation prevalence remained more than three times higher than urban prevalence throughout the period. Although the ratio showed a consistent downward trend, the magnitude of reduction was limited, reflecting persistent relative inequality.

The attributable inequality measures also showed gradual improvement over time. The population attributable fraction (PAF) increased in absolute terms from −62.62% in 2000 to −48.85% in 2024, suggesting a reduction in the proportion of national open defecation attributable to rural–urban differences, although substantial inequality remained. Similarly, the population attributable risk (PAR) declined in absolute magnitude from −12.72 to −8.69 percentage points, indicating a slow reduction in the excess national burden associated with residence-based disparities.

Overall, all four-inequality metrics consistently indicate sustained rural–urban disparities in open defecation over the study period. While all measures show gradual improvement, the pace of change is modest and insufficient to indicate meaningful convergence between rural and urban populations.

### Joint Point Regression Analysis

Between 2000 and 2024, national open defecation in Ghana showed a statistically significant declining trend with one identified joinpoint in 2016 (95% CI: 2015–2017). From 2000 to 2016, which broadly corresponds to the Millennium Development Goal (MDG) period and the immediate transition phase, prevalence declined at an annual percentage change (APC) of −0.5674% (95% CI: −0.5703 to −0.5650; p < 0.001). This was followed by a slightly slower decline of −0.52% per year (95% CI: −0.53 to −0.51; p < 0.001) from 2016 to 2024, corresponding to the Sustainable Development Goal (SDG) era. The average annual percentage change (AAPC) over the full period was −0.55%, indicating a consistent and statistically significant downward trend.

In rural areas, no statistically significant joinpoints were identified over the study period, indicating a stable linear trend across both the MDG and SDG periods. The APC for rural areas was +0.07% per year (95% CI: +0.07 to +0.07; p < 0.001), and the AAPC was identical, reflecting a persistent but very small upward trajectory without detectable structural change over time.

In urban areas, two statistically significant joinpoints were identified in 2008 (95% CI: 2003–2011) and 2015 (95% CI: 2012–2020), resulting in three distinct trend segments spanning both MDG and SDG periods. From 2000 to 2008 (MDG period), prevalence increased at an APC of +0.80% (95% CI: +0.79 to +0.84; p < 0.001). This was followed by a slightly lower increase of +0.76% per year (95% CI: +0.74 to +0.78; p < 0.001) from 2008 to 2015 (late MDG transition period), and a further modest deceleration to +0.72% per year (95% CI: +0.69 to +0.73; p < 0.001) from 2015 to 2024 (SDG period). The AAPC over the full period was +0.76% (95% CI: +0.75 to +0.76; p < 0.001), indicating a sustained upward trend with gradual slowing over time. These trends are shown in Table 3 and figure 2. The results indicate heterogeneous temporal patterns across residence groups across both MDG and SDG eras. While the national trend shows a statistically significant decline, rural areas exhibit a persistent upward trajectory without structural change, and urban areas show a sustained increase with gradual deceleration across successive policy eras.

**Figure 1:**
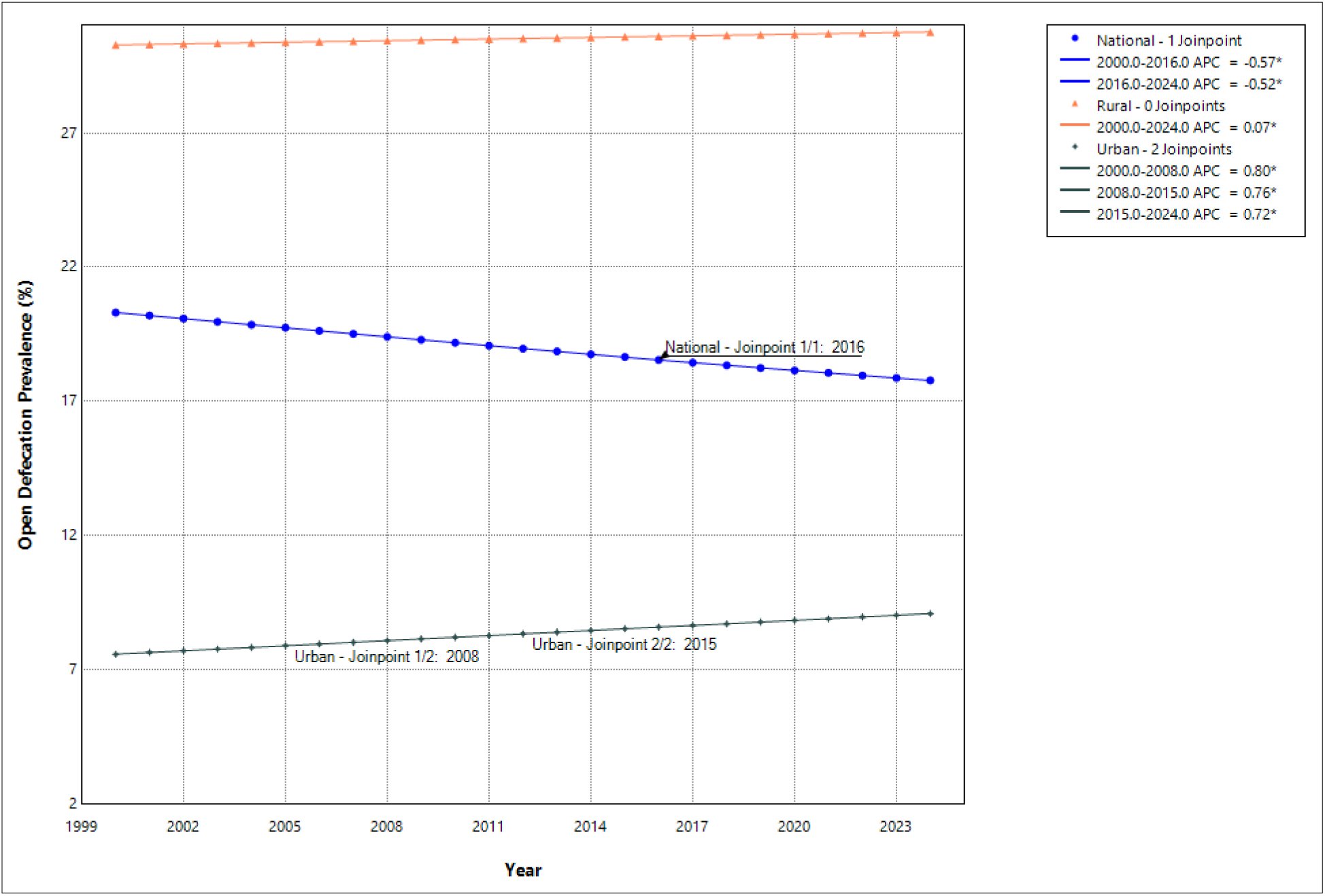
Joinpoint regression of national, rural, and urban open defecation prevalence trends in Ghana, 2000–2024

**Figure 2:**
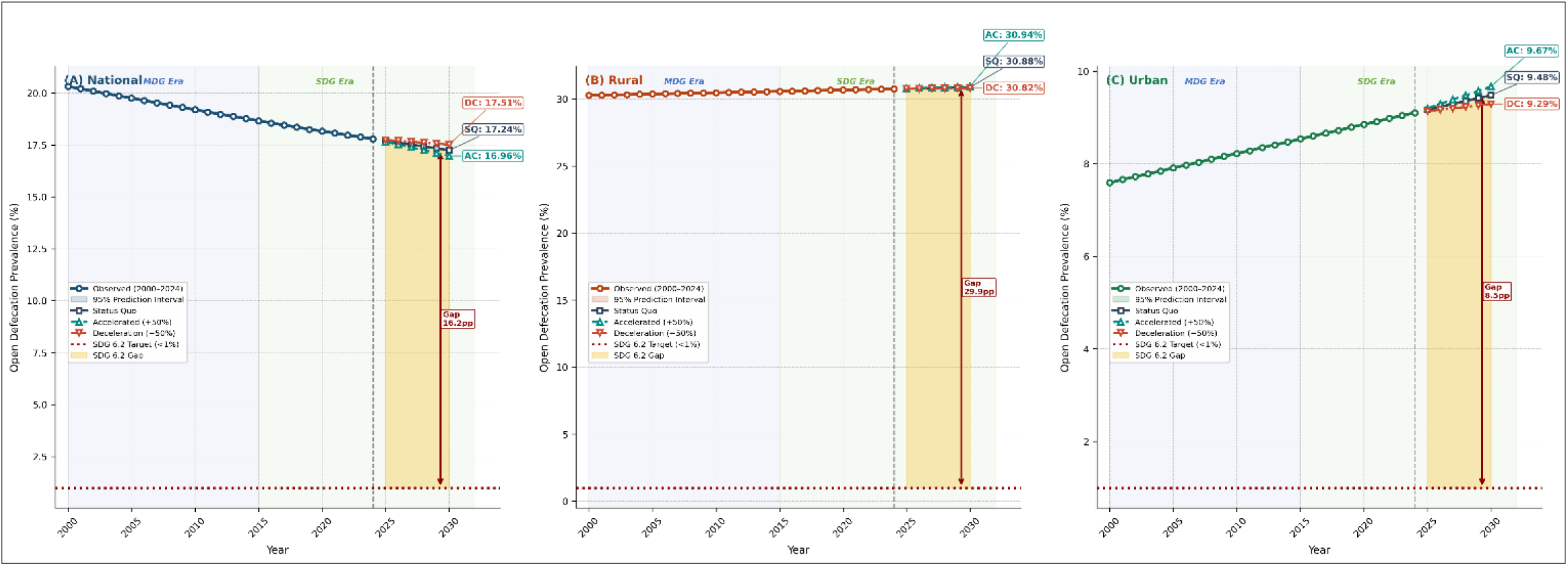
Open Defecation Prevalence in Ghana: Observed Trends and Scenario-Based Projections to 2030-National, Rural, and Urban (ARIMA Model, 2000-2024 Baseline)

**Table 3:**
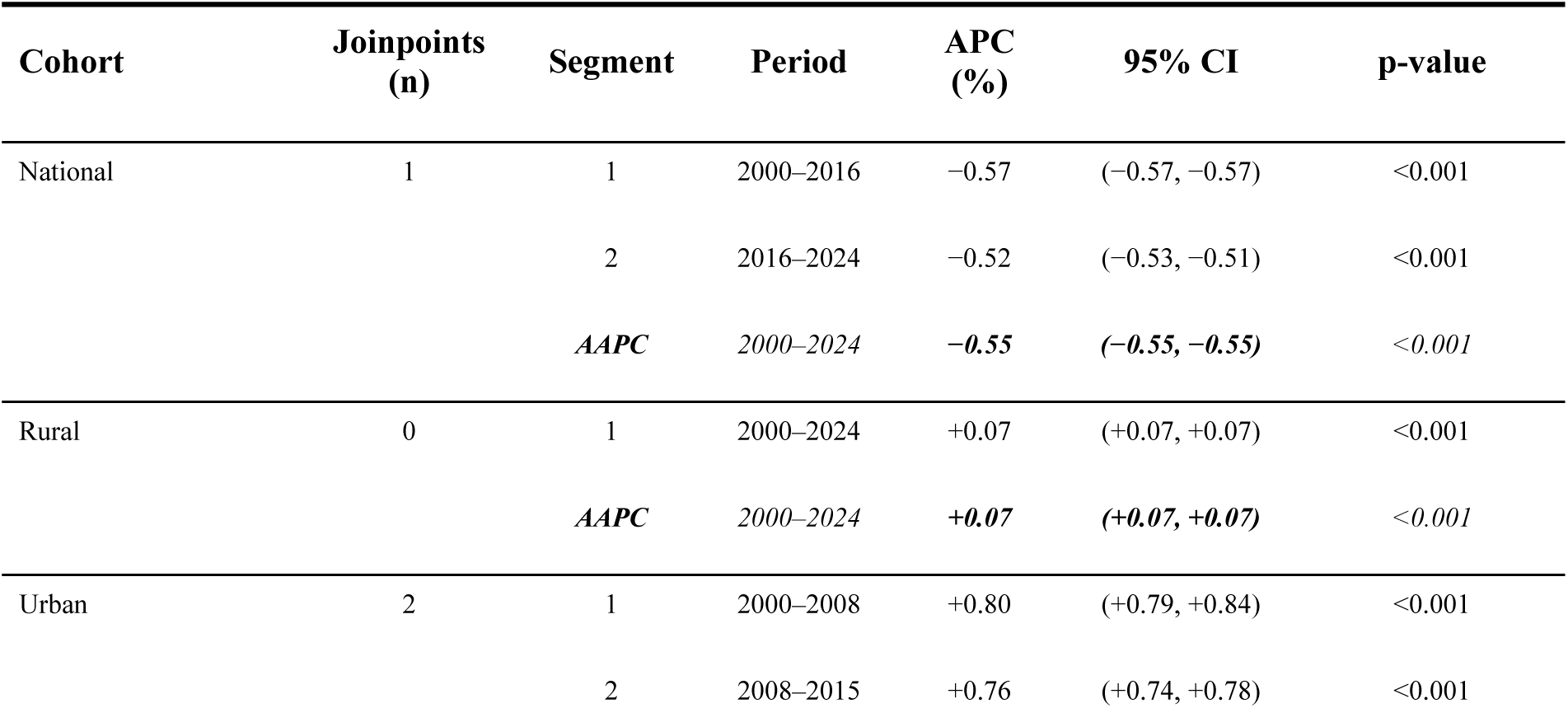
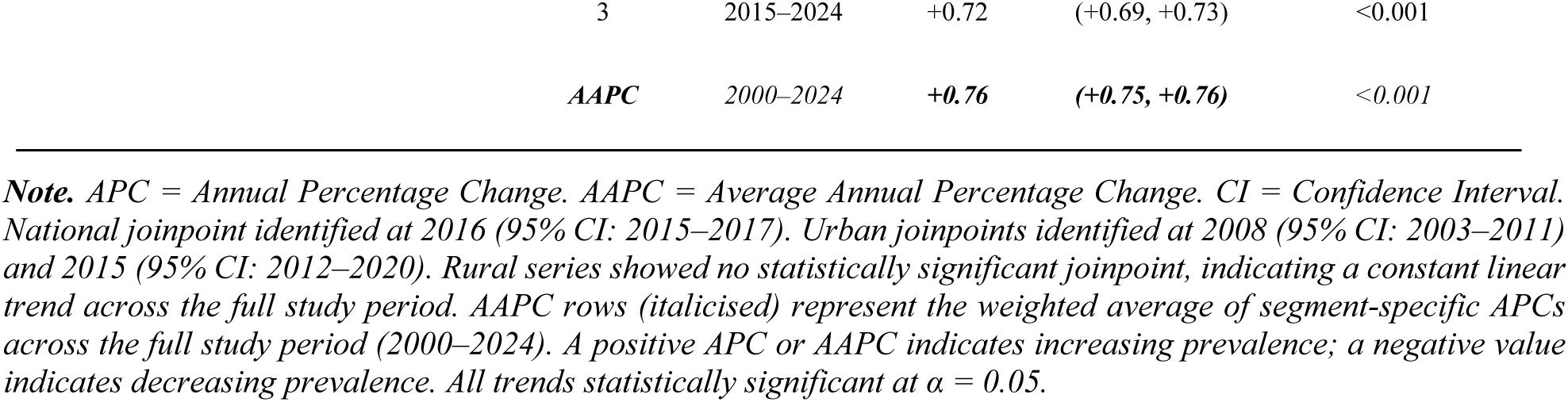
Joinpoint Regression Analysis of Open Defecation Prevalence Trends by Residence Group, Ghana, 2000–2024.

### Forecast Analysis and Projections to 2030

The ARIMA forecasting analysis showed clear subgroup differences in the trajectory of open defecation prevalence in Ghana from 2025 to 2030. The national series was stationary, while the rural and urban series were non-stationary and required differencing. Across all three series, model fit and validation performance were strong, with low forecast errors and residual diagnostics indicating no meaningful autocorrelation.

For the national series, the best-fitting model for both validation and the full sample was ARIMA (0,2,2). The hold-out validation showed very high accuracy, with MAE of 0.0241 percentage points, RMSE of 0.0257, and MAPE of 0.1346%. Residual diagnostics were satisfactory, and the final model had an AIC of-169.834. The forecast projected a gradual decline in national open defecation prevalence from 17.70% in 2025 to 17.24% in 2030 under the status quo scenario. Under the accelerated scenario, prevalence declined slightly faster, reaching 16.96% in 2030, while the deceleration scenario ended at 17.51%. Although the national trend was downward, none of the scenarios approached the SDG 6.2 target of less than 1.0%, leaving a gap of 15.96 to 16.51 percentage points in 2030.

For the rural series, the best-fitting validation model was ARIMA (2,2,1), while the final full-series model was ARIMA (0,2,0), with a very low AIC of-502.182. Validation performance was excellent, with MAE, RMSE, and MAPE all equal to 0.0000, and residual diagnostics showed no evidence of autocorrelation. However, unlike the national series, the rural forecasts showed a slight upward trend over time. Status quo prevalence increased from 30.78% in 2025 to 30.88% in 2030. The accelerated scenario produced even higher values, reaching 30.94% by 2030, while the deceleration scenario ended at 30.82%. This occurs because the accelerated scenario amplifies the direction of the projected trend; since the rural series is rising, the accelerated path rises faster, whereas the deceleration path rises more slowly. Despite the high model accuracy, all rural scenarios remained far above the SDG threshold, with a 2030 gap of about 29.82 to 29.94 percentage points.

For the urban series, the best-fitting model for both validation and the full sample was ARIMA (3,2,1), with a final AIC of-185.427. Validation results also indicated excellent fit, with MAE of 0.0014, RMSE of 0.0023, and MAPE of 0.0156%, and residuals were free of meaningful autocorrelation. The urban forecasts likewise showed a gradual increase over time. Status quo prevalence was projected to rise from 9.17% in 2025 to 9.48% in 2030. The accelerated scenario increased more rapidly to 9.67%, while the deceleration scenario rose more slowly to 9.29%. As with the rural series, the accelerated values are higher because the fitted trajectory is upward and the scenario rules magnify or dampen that increase. Even though urban prevalence was lower than the rural and national estimates, it still remained far above the SDG 6.2 target, with a 2030 gap of 8.29 to 8.67 percentage points, see Figure 2, and Table 4.

**Table 4:**
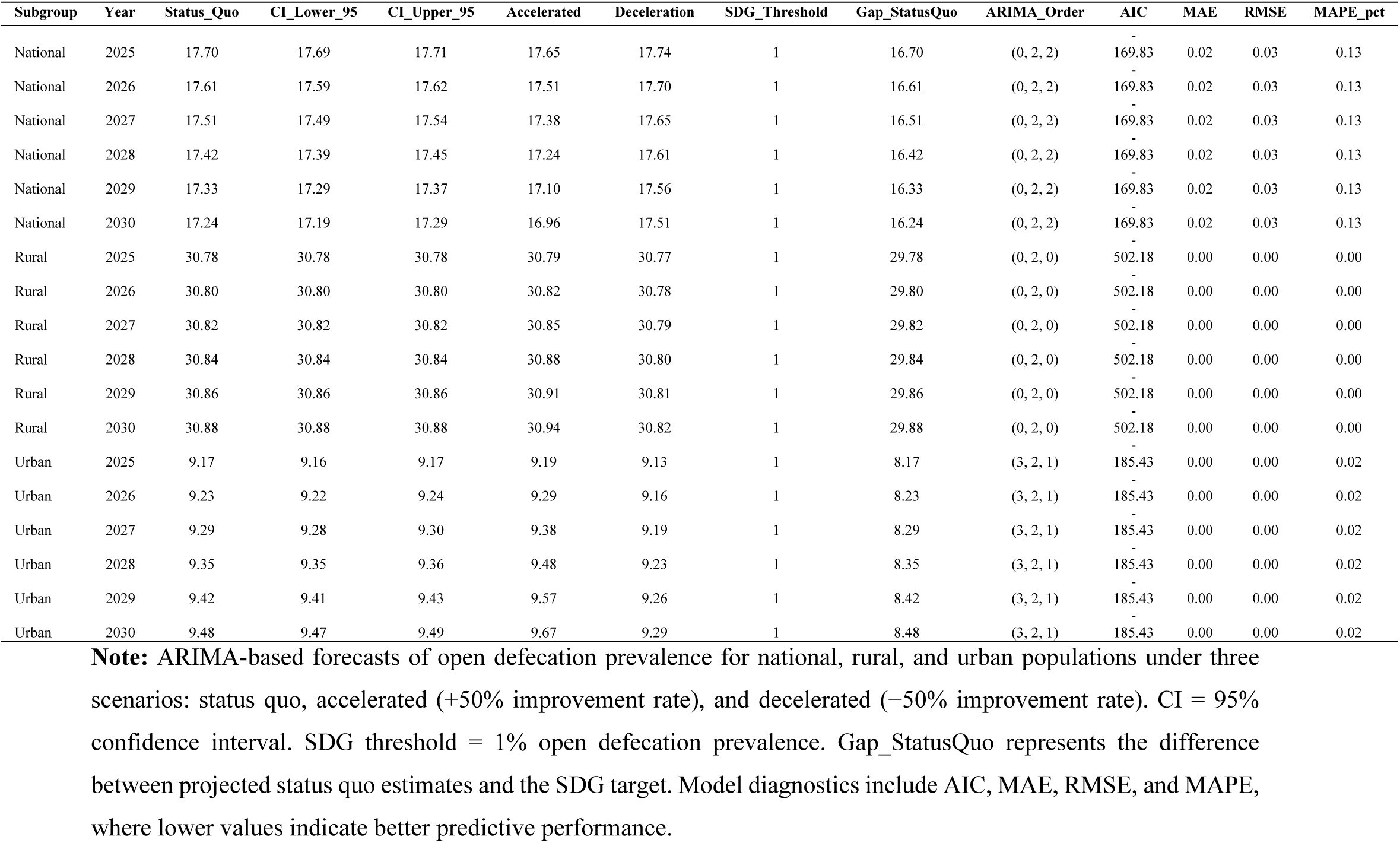
National, rural, and urban open defecation forecasts with ARIMA diagnostics and 2030 SDG gap.

Overall, the forecasts suggest that Ghana is unlikely to achieve the SDG 6.2 open defecation target by 2030 under any of the modeled scenarios. The national series indicates a modest decline, but the rural and urban series are projected to rise, showing that progress is not uniform across residence groups. The rural population continues to carry the greatest burden, while the urban series, though lower, also remains well above the target. These findings imply that stronger and more targeted interventions are needed, especially in rural areas, if open defecation is to be reduced meaningfully over time.

## Discussion

This study analysed trends, rural–urban inequalities, and projections of open defecation in Ghana from 2000 to 2024 using WHO standardised inequality measures, joinpoint regression, and ARIMA forecasting. Three principal findings emerge. First, national open defecation prevalence declined modestly. Second, substantial rural–urban inequalities persisted despite slight improvements in absolute and relative measures. Third, projections indicate that Ghana is unlikely to achieve SDG 6.2 by 2030. These findings demonstrate that while incremental progress has occurred, its pace and distribution remain insufficient.

The modest national decline (from 20.31% in 2000 to 17.79% in 2024), averaging less than 1% per year, is consistent with regional evidence that sanitation improvements across Sub-Saharan Africa have lagged population growth and urban expansion, leaving many countries off track toward SDG targets [14,26]. Persistent structural barriers, including poverty, uneven sanitation financing, limited household affordability of improved facilities, and implementation weaknesses in community sanitation programmes, plausibly account for Ghana’s constrained progress [33,34]. Globally, WHO/UNICEF monitoring suggests that safely managed sanitation is progressing too slowly to achieve universal coverage by 2030 [11,35], highlighting the compounding roles of governance failures, weak political commitment, and persistent inequities in financing.

However, a critical interpretive caveat must be acknowledged. The observed national decline occurred despite rural and urban open defecation prevalence both increasing over the same period. This apparent paradox is consistent with Simpson’s Paradox, whereby changes in population composition can produce aggregate trends that differ from, or even oppose, trends observed within constituent subgroups [36]. This suggests that a substantial portion of Ghana’s apparent national progress may reflect demographic compositional change driven by urbanisation rather than genuine sanitation improvement [36]. This finding has important implications for how national-level sanitation data are interpreted and reported, and underscores the necessity of routinely disaggregating estimates by residence and other equity dimensions.

The persistence of rural–urban inequalities is among the most consequential findings. Across all four WHO measures, rural populations consistently carried a higher open defecation burden, remaining more than three times higher than urban prevalence by 2024. This aligns with evidence from Sub-Saharan Africa and Ghana, where open defecation disproportionately affects rural and socioeconomically disadvantaged populations, particularly those in lower wealth quintiles, and is driven by lower educational attainment, geographic isolation, inadequate sanitation infrastructure, and weak enabling systems [17,29,37–40]. Ghana’s rural–urban disparity mirrors patterns observed in India, where aggregate national gains masked persistent deprivation among remote populations [37 [41], highlighting a broader failure of equity-sensitive sanitation monitoring. These inequalities reflect interacting infrastructural, socioeconomic, financing, institutional, geographic, and behavioural barriers; socially embedded sanitation norms may further weaken infrastructure-focused interventions without sustained behaviour change strategies [18,40,42,43].

Attributable inequality measures provide further policy insight. Although PAR and PAF improved modestly (from −12.72 to −8.69 percentage points and from −62.62% to −48.85%, respectively), a substantial proportion of the national burden remains attributable to rural disadvantage, suggesting that equity-targeted rural interventions, rather than aggregate national programmes, may be the most efficient lever for reducing overall prevalence [51 (19)]. Equalising rural open defecation to urban levels could reduce the national burden by nearly half, highlighting a fundamental shortfall in Ghana’s sanitation policy reach and equity orientation [44,45].

Joinpoint regression identified a single national inflection at 2016 (95% CI: 2015–2017), coinciding with the MDG-to-SDG transition. However, the annual rate of decline slowed slightly during the SDG era (APC: −0.52% per year) compared with the preceding MDG period (−0.57% per year), suggesting challenges in sustaining earlier gains. Notably, no rural joinpoints were identified across the 25-year period, with prevalence following a near-linear upward trajectory (APC: +0.07% per year), indicating that neither MDG-era nor SDG-era programmes, including Ghana’s CLTS implementation from 2008 onward, generated meaningful structural shifts in rural sanitation behaviour or coverage. This absence of inflection in rural trends is consistent with documented evidence of sanitation slippage and weak post-triggering sustainability in Ghana’s CLTS programme [20,46], and suggests that programmatic gains were insufficient to overcome the structural and behavioural barriers driving rural open defecation.

Urban prevalence increased throughout the study period, although the rate of increase slowed gradually across successive periods (APC: +0.80%, +0.76%, and +0.72%), consistent with rapid urbanisation, expansion of informal settlements, and infrastructure development failing to keep pace with population growth [47,48]. Importantly, urban residence does not guarantee equitable sanitation access, particularly for residents of informal settlements who lack tenure security or financial capacity to afford improved facilities [49]. The sustained urban upward trend across 25 years, despite economic growth and urbanisation, challenges the common assumption that urbanisation inherently drives sanitation improvement and warrants dedicated policy attention to urban sanitation equity. Future analyses should disaggregate urban populations by settlement type and socioeconomic status to better characterise within-urban heterogeneity.

The broader public health consequences of persistent open defecation extend well beyond diarrhoeal disease. Open defecation facilitates the transmission of soil-transmitted helminths and enteropathogens, contributing to intestinal infection, impaired nutrient absorption, and poor child growth [50]. Evidence from major WASH trials in Kenya, Bangladesh, and Zimbabwe indicates that sanitation and hygiene interventions can reduce some faecal exposure pathways, but have shown limited and context-dependent effects on linear growth and stunting [51,52]. In Ghana, childhood stunting remains around 18% nationally and is highest in northern regions, where sanitation deprivation and open defecation are also concentrated [53]. Environmental enteric dysfunction (EED), helminth co-infection, and diarrhoeal disease may therefore interact to reduce nutrient bioavailability and energy reserves [54,55]. Poor sanitation may further compound vulnerability through EED-mediated impairment of mucosal immunity, though the sanitation–EED–vaccine efficacy pathway should be considered biologically plausible rather than conclusive given mixed trial evidence [54,56–58]. Contaminated irrigation water and pathogen detection in vegetables additionally highlight open defecation and poor faecal waste management as food safety hazards [59].

Gendered dimensions further compound the sanitation burden. Although not explored in this study, evidence indicates that women and girls face increased exposure to gender-based violence when defecating in the open, while poor sanitation elevates urogenital infection risk and adversely affects reproductive health and pregnancy outcomes [60,61]. Inadequate menstrual hygiene management in schools and workplaces contributes to absenteeism and school dropout, with long-term consequences for female human capital, economic participation, and intergenerational poverty [62]. Rural women, who bear the greatest burden of open defecation, remain structurally disempowered to demand infrastructural remedies, highlighting the intersection of sanitation deprivation, gender inequality, and social marginalisation [63,64].

Open defecation and inadequate sanitation may also contribute to antimicrobial resistance (AMR) by increasing faecal contamination of community environments. Evidence indicates that poor sanitation and limited wastewater treatment increase environmental loads of antibiotic-resistant bacteria and resistance genes, creating conditions for environmental persistence and horizontal gene exchange [65,66]. In Ghana specifically, ESBL-producing E. coli has been identified in river waters in Accra and Tamale, and genomic evidence has demonstrated diverse beta-lactamase genes in multidrug-resistant E. coli from environmental water sources [66–71]. Although Ghana’s AMR policy architecture acknowledges sanitation and hygiene within a One Health response, peer-reviewed analyses suggest that implementation has remained more strongly centred on antimicrobial stewardship, surveillance, and coordination than on structural environmental determinants [72,73], underscoring the wider consequences of failing to achieve sanitation-related SDG targets.

Forecast analyses reinforce the urgency of intervention. Under the status quo, national prevalence was projected at 17.24% by 2030, while rural prevalence reached 30.88%, nearly thirty percentage points above the SDG 6.2 threshold. The ARIMA-projected upward rural trend is consistent with population growth outpacing facility construction and with sanitation slippage following CLTS triggering, both of which are documented in Ghana and across Sub-Saharan Africa [20,46]. Parallels with India’s post-Swachh Bharat verification surveys highlight the broader risk of decoupling programmatic certification from sustained behavioural and infrastructural outcomes [74,75], and Ghana’s quantitative forecasts add longitudinal precision to this pattern.

### Policy Implications

Policy implications are clear. Ghana requires equity-focused sanitation strategies that explicitly target rural and underserved populations, where open defecation remained persistently concentrated throughout the study period. The CLTS programme requires stronger and more sustained investment, rigorous post-triggering monitoring, and structural integration with poverty reduction and social protection initiatives to minimise sanitation slippage and improve long-term sustainability. Behavioural and social norm interventions must complement infrastructure provision, particularly in rural communities where open defecation remains socially embedded. National sanitation monitoring systems should routinely incorporate disaggregated inequality indicators, including residence, wealth, and region, alongside aggregate national estimates to prevent persistent rural deprivation from being obscured by demographic compositional shifts.

The findings further suggest that sanitation improvement is not solely a behavioural challenge but fundamentally a structural one. Despite two decades spanning both MDG and SDG eras, rural open defecation trends showed no meaningful structural inflection, indicating that interventions implemented without broader enabling conditions, including household affordability, sustained infrastructure investment, and institutional capacity, are unlikely to achieve durable impact. Future sanitation policy must therefore integrate behaviour change approaches within a long-term structural investment framework targeting the root determinants of rural sanitation deprivation.

### Strengths and Limitations

Strengths of this study include the integration of four WHO-standardised inequality measures, joinpoint regression, and ARIMA forecasting within a unified analytical framework spanning 25 years across both MDG and SDG eras. The use of internationally comparable WHO HEAT metrics enabled simultaneous assessment of absolute and relative rural–urban inequalities using nationally representative data [25]. The hold-out validation approach for ARIMA models, yielding MAPE values well below 1% across all series, further strengthens confidence in the forecast estimates. Combining historical trend analysis with forward projections strengthened the study’s policy relevance by providing evidence not only on past and current sanitation patterns but also on Ghana’s likely trajectory toward SDG 6.2.

However, several limitations warrant consideration. The study relied on secondary aggregated data and could not assess household-level determinants or causal pathways. Analyses were restricted to place of residence and did not examine other important inequality dimensions such as wealth, education, or subnational region. The study also did not account for potential disruptions to sanitation behaviours and data collection arising from the COVID-19 pandemic during 2020–2022, which may have affected observed trends in that period. Finally, the Simpson’s Paradox dynamic identified in this study whereby national prevalence declined despite worsening rural and urban subgroup trends, highlights the risk of relying on aggregate national estimates for sanitation monitoring without simultaneous disaggregation [36,76]. Future studies incorporating, within-urban disaggregation, and sensitivity analyses would substantially strengthen analytical robustness.

## Conclusion

Despite modest national progress, rural–urban sanitation inequalities remain deeply entrenched, and Ghana is unlikely to achieve SDG 6.2 under any of the modelled trajectories. The apparent national decline is substantially explained by demographic compositional change rather than genuine sanitation improvement across population subgroups, as both rural and urban open defecation prevalence increased over the study period. Persistent open defecation continues to contribute to infectious disease transmission, child undernutrition, food contamination, gendered vulnerability, and antimicrobial resistance. Accelerated, equity-focused interventions targeting the structural determinants of rural sanitation deprivation, including sustained infrastructure investment, post-triggering CLTS monitoring, and integration with poverty reduction, will be critical to closing the rural–urban gap and improving Ghana’s trajectory toward SDG 6.2. The analytical framework applied in this study provides a replicable model for longitudinal sanitation inequality assessment across Sub-Saharan Africa.

## Abbreviations

AAPC: Average Annual Percentage Change
ADF: Augmented Dickey-Fuller
AIC: Akaike Information Criterion
AMR: Antimicrobial Resistance
APC: Annual Percentage Change
ARIMA: Autoregressive Integrated Moving Average
BIC: Bayesian Information Criterion
CI: Confidence Interval
CLTS: Community-Led Total Sanitation
CWSA: Community Water and Sanitation Agency
DHS: Demographic and Health Surveys
EED: Environmental Enteric Dysfunction
ESBL: Extended-Spectrum Beta-Lactamase
HEAT: Health Equity Assessment Toolkit
JMP: Joint Monitoring Programme
MAE: Mean Absolute Error
MAPE: Mean Absolute Percentage Error
MDG: Millennium Development Goal
MICS: Multiple Indicator Cluster Surveys
ODF: Open Defecation Free
PAF: Population Attributable Fraction
PAR: Population Attributable Risk
RMSE: Root Mean Square Error
SDG: Sustainable Development Goal
UNICEF: United Nations Children’s Fund
WASH: Water, Sanitation and Hygiene
WHO: World Health Organization

## Data Availability

No new data were generated in this study. The data underlying the findings are publicly available from the World Health Organization Health Equity Assessment Toolkit (WHO HEAT) and the WHO Health Inequality Monitor database. The datasets analysed during the current study can be accessed without restriction at https://www.who.int/data/inequality-monitor/data. All relevant data are contained within the paper and its Supporting Information files.

## Acknowledgements

We thank the World Health Organization (WHO) and the United Nations Children’s Fund (UNICEF) for making the WHO Health Equity Assessment Toolkit (HEAT) platform and its underlying datasets publicly available, which formed the sole data source for this analysis.

## Authors’ contributions

AI and MIN conceived the study. AI and MN wrote the methods section and performed the data analysis. AI and MIN, were responsible for the initial draft of the manuscript. AS critically reviewed the initial draft. All the authors reviewed and approved the final version of the manuscript.

## Ethics Declarations

### Ethics approval and consent to participate

Not applicable. This study used publicly available, de-identified, aggregated secondary data obtained from the WHO Health Equity Assessment Toolkit (HEAT) platform. No primary data collection was undertaken and no human participants were involved. Ethical approval was therefore not required under institutional guidelines.

### Consent for publication

Not applicable.

### Availability of data and materials

All data used in this study are publicly available through the World Health Organization Health Equity Assessment Toolkit (WHO HEAT) platform. The underlying datasets are accessible from the WHO Inequality Monitor database at https://www.who.int/data/inequality-monitor/data. No restrictions apply to the availability of these data.

## Supplementary Information

**Supplementary Figure S1.**
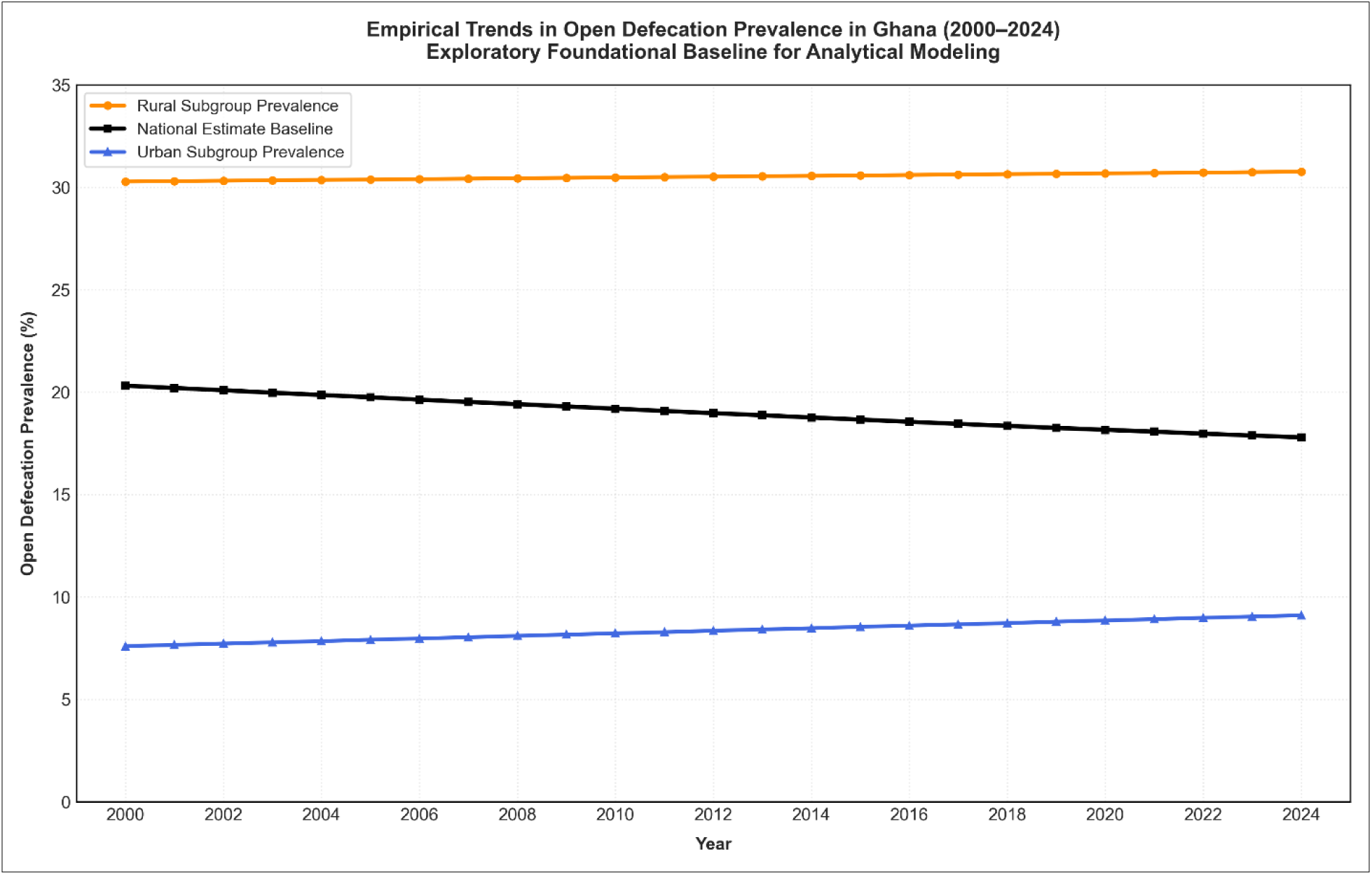
Empirical Trends in Open Defecation Prevalence in Ghana, 2000–2024: Exploratory Baseline for Analytical Modeling

**Supplementary Table S2.** Joinpoint regression model outputs for national open defecation prevalence trends in Ghana, 2000–2024.

**Supplementary Table S3.** Joinpoint regression model outputs for rural open defecation prevalence trends in Ghana, 2000–2024.

**Supplementary Table S4.** Joinpoint regression model outputs for urban open defecation prevalence trends in Ghana, 2000–2024.

**Supplementary Figure 2:**
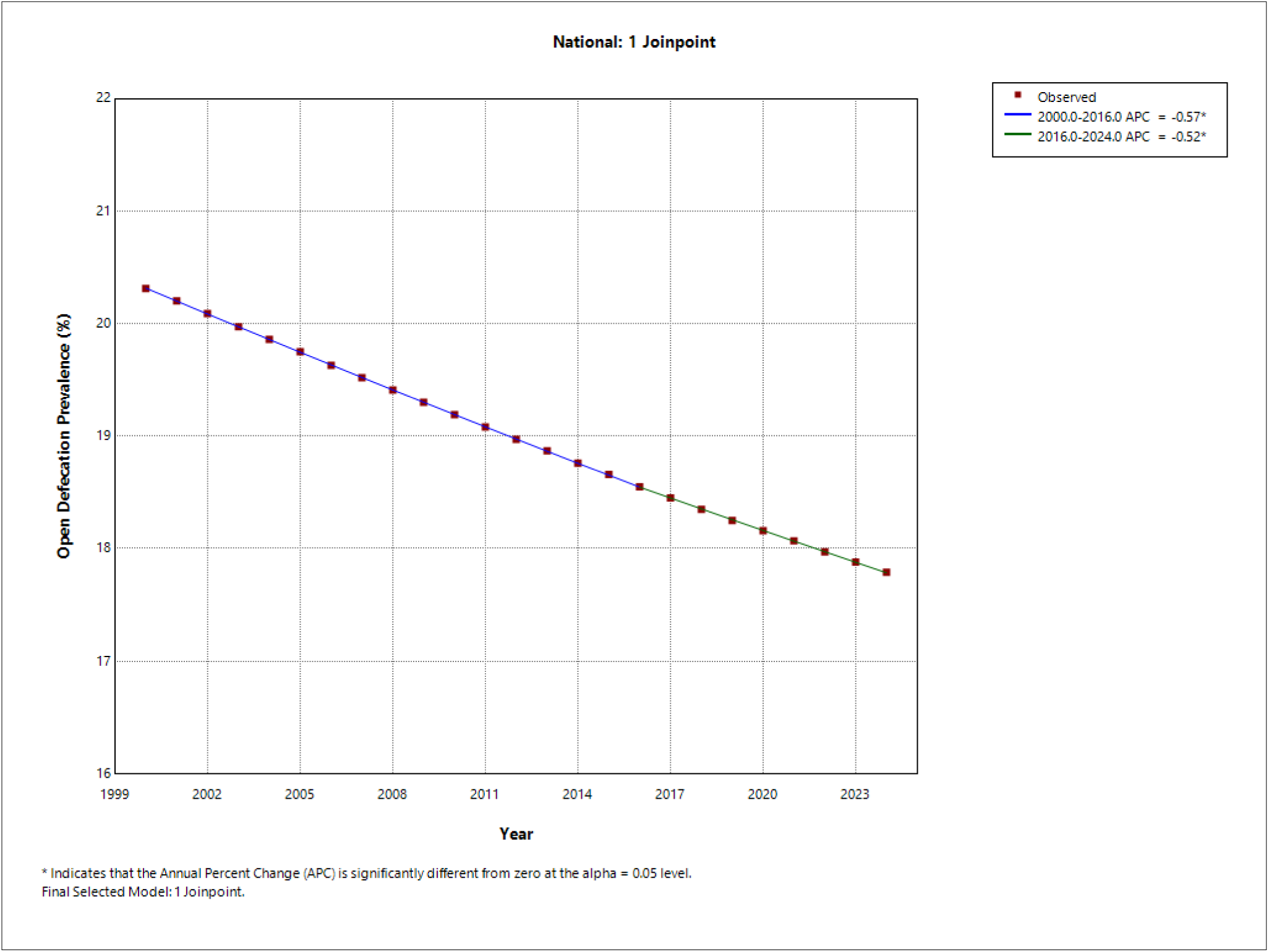
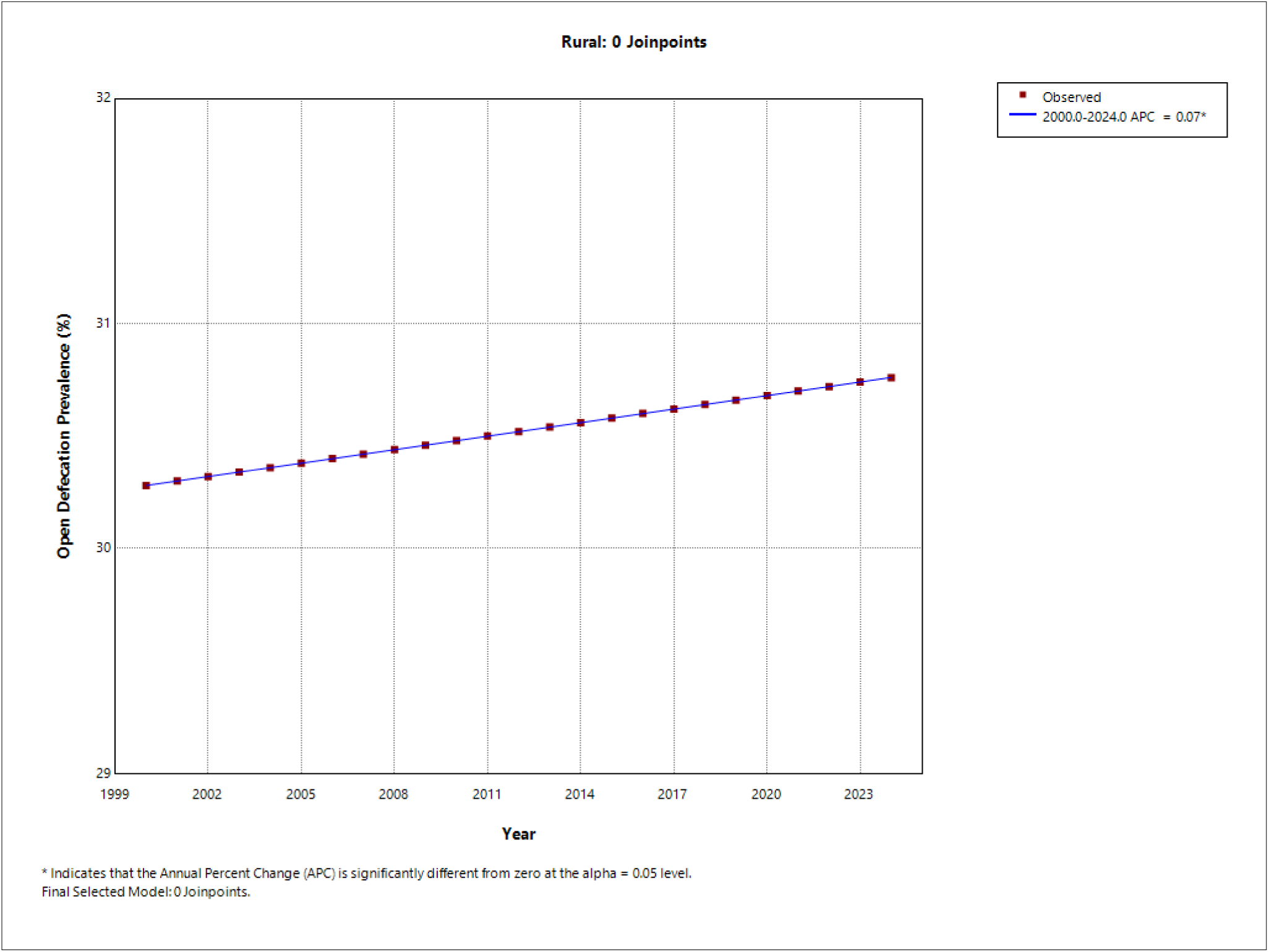
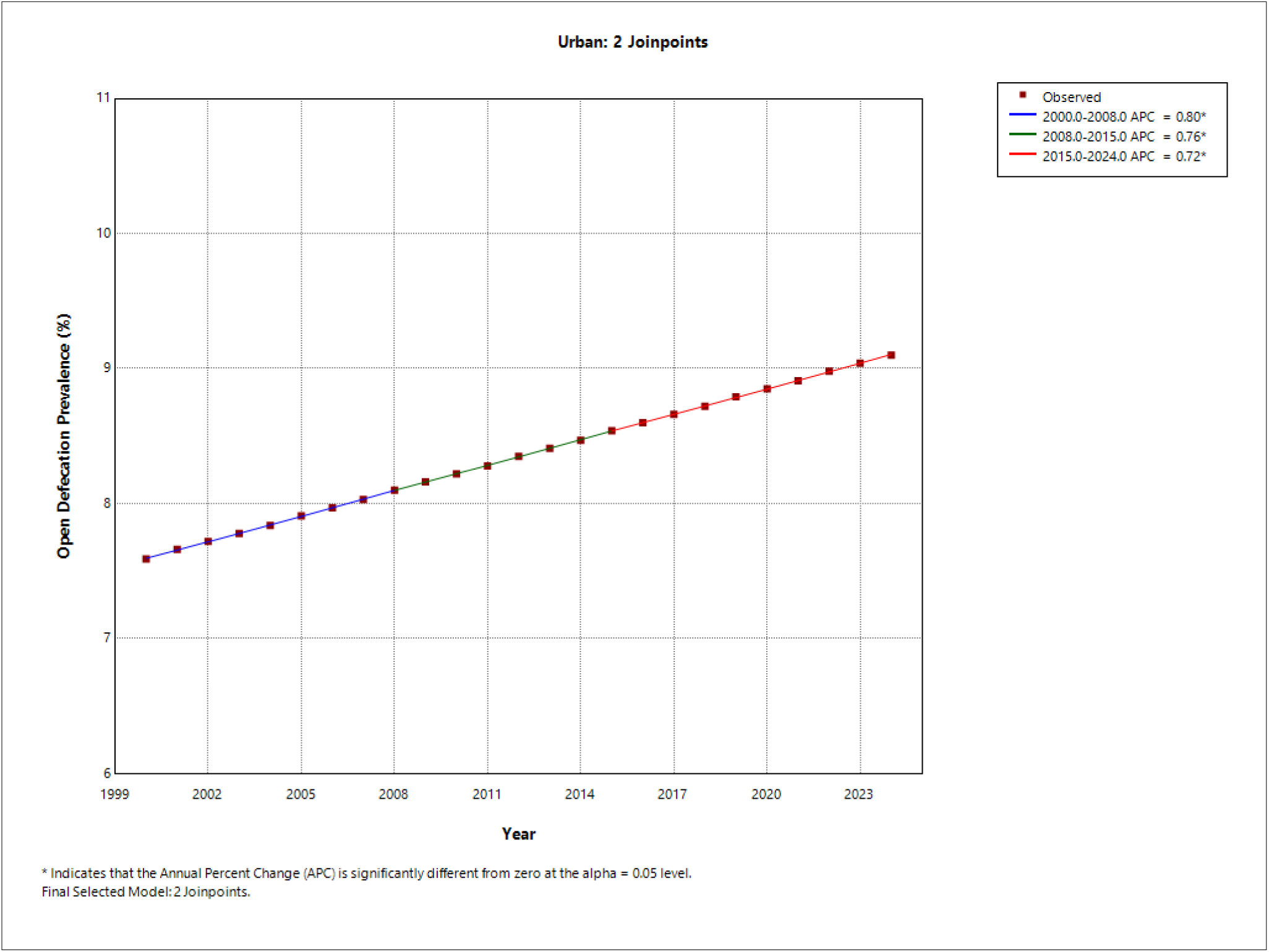
Ghana Open Defecation Prevalence — Observed Trends and Status Quo Projections to 2030 by Residence Group with SDG 6.2 Gap.

**Supplementary Figure 3:**
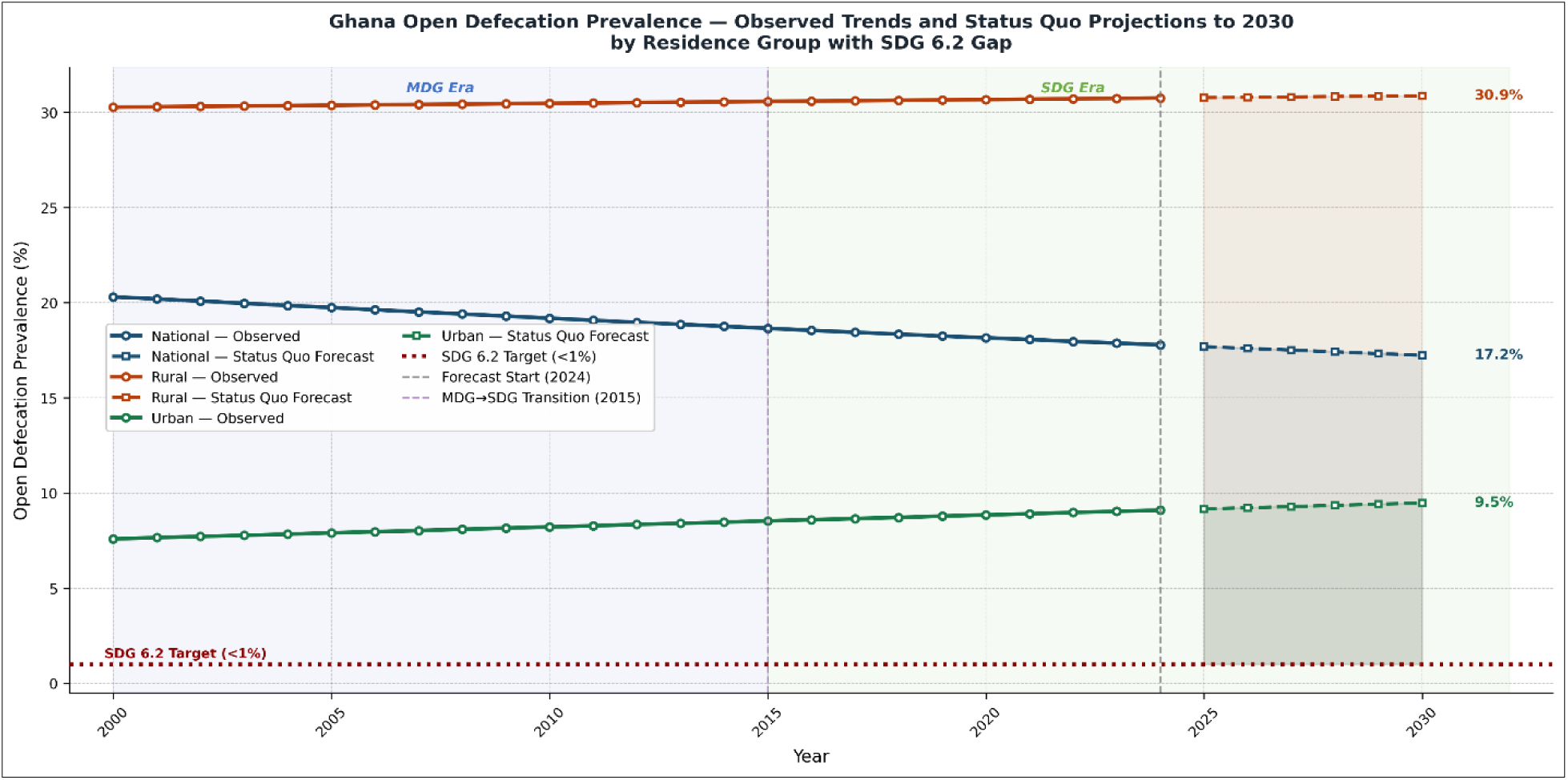
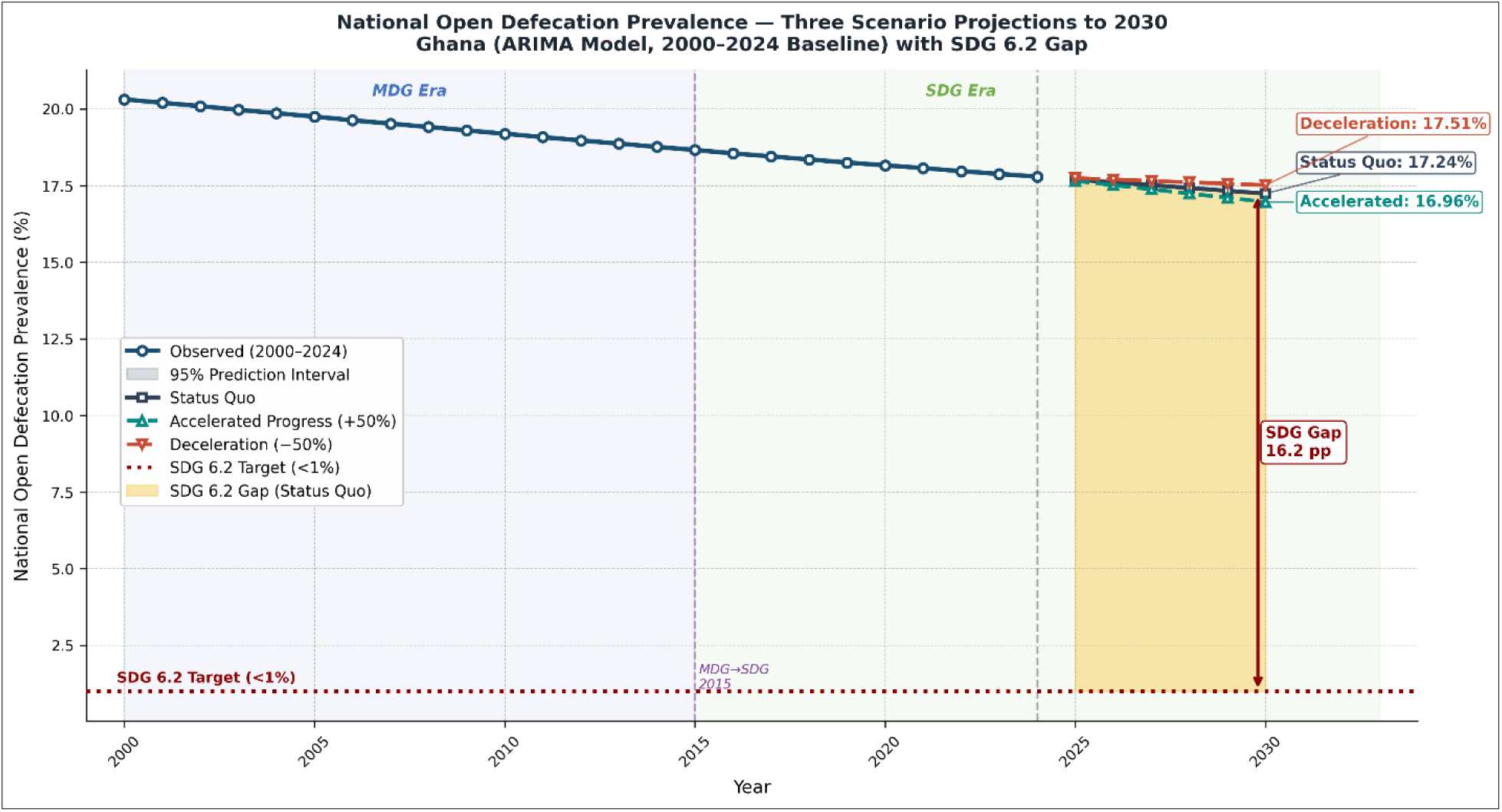
National Open Defecation Prevalence —Three Scenario Projections to 2030, Ghana (ARIMA Model, 2000–2024 Baseline) with SDG 6.2 Gap.

